# ECG-derived age deviation predicts cardiovascular diseases across lead configurations and cohorts

**DOI:** 10.64898/2026.06.05.26354974

**Authors:** Deniz Aydogdu, Farieda Gaber, Arash Sorooshmehr, Altuna Akalin

## Abstract

Cardiovascular diseases (CVDs) remain the primary global health burden, motivating the search for robust, non-invasive risk biomarkers. We harness a foundation model pretrained on over 10 million recordings, to evaluate ECG-derived age deviation as a cross-cohort biomarker of CVD burden. A predictive model, trained exclusively on healthy subjects, achieved accurate age prediction. Diseased subjects exhibited significant positive age acceleration across multiple categories, with structural and ischemic heart diseases showing the largest effects. External validation in a hospital-based cohort (n=160,493) confirmed that age acceleration independently predicts all-cause mortality, with the strongest prognostic value in patients under 65 years. Furthermore, we demonstrated that disease discrimination and mortality prediction are preserved across 6-lead and single-lead configurations, supporting potential deployment in wearable or mobile devices. Our analysis also revealed a striking morphological confound from the complete left bundle branch block, leading us to propose absolute age deviation as a more robust, universal risk marker. These findings establish ECG-derived biological age deviation as a highly generalizable and clinically actionable biomarker for assessing cardiovascular risk. We have also developed a web application at https://bioinformatics.mdc-berlin.de/ECGage that allows users to easily test our framework.

## Introduction

Cardiovascular diseases (CVDs) are the leading cause of death worldwide, responsible for 19.2 million deaths in 2023, approximately one-third of all global mortality according to the latest Global Burden of Disease report^1^. The global impact of CVD is expected to remain substantial, driven in part by population growth, population aging, and the rising prevalence of metabolic risk factors across 204 countries and territories ^1^. Population aging is particularly relevant because age is one of the strongest cardiovascular risk factors. However, in clinical practice, age is usually represented by chronological age (CA), although individuals of the same age can vary substantially in their physiological states^1–3^. Biological age (BA) therefore aims to capture this variation by estimating physiological status rather than the mere passage of time. The discrepancy between CA and BA, termed age deviation, represents a hidden window into cardiovascular risk that traditional risk assessments, which often rely on CA, overlook^4^.

The electrocardiogram (ECG) has emerged as a highly accessible source of biomarkers for detecting cardiovascular diseases with prognostic relevance^5,6^. Deep learning models can be trained to predict CA directly from ECG signals. The difference between ECG-derived predicted age and CA, known as age deviation, is associated with mortality and adverse cardiovascular outcomes^7,8,9^. These findings suggest that the predicted age captures critical pathophysiological signals beyond CA.

Despite advances in deep learning-based ECG-derived age prediction, most studies utilize institution-specific and task-specific models, which inherently limits their reproducibility and generalizability. Recently, the paradigm in medical AI has shifted toward foundation models, which offer robust, broadly transferable representations from large-scale datasets that address the limitations of narrow, task-specific training. At the same time, the growing interest in wearable and portable ECG monitoring has motivated frameworks such as PROPHECG-Age^10^, which explore age estimation from single-lead ECG configurations.However, even with powerful representations of foundation models, several critical knowledge gaps remain in the clinical application of foundation models for ECG-derived age deviation. First, the disease-specific patterns of age deviation, specifically which conditions drive positive versus negative age deviation remain incompletely characterized. Second, conduction abnormalities can fundamentally alter ECG morphology in ways that may systematically confound embedding-based age prediction^11^, yet this phenomenon has not been systematically isolated or examined with models capable of capturing diverse ECG morphologies and rhythms^12^. Third, as cardiovascular monitoring increasingly shifts towards wearables devices, the feasibility of measuring biological age from reduced-lead configurations remains a significant gap^13,14^. Lastly, the clinical utility of these AI-derived biomarkers requires external validation against hard clinical outcomes, such as mortality^15,16^.

To address these gaps, we used ECGFounder^1,30^, a publicly available foundation model pretrained on over 10 million recordings, to evaluate ECG-derived age deviation as a robust cardiovascular disease biomarker (The workflow visualized in Figure 1). By training an age prediction model exclusively on healthy subjects from the PTB-XL dataset^17^, we aimed to systematically examine age deviation across diverse disease categories and isolate disease-specific deviations from natural senescence. We specifically address and isolate conduction abnormalities as a major source of morphological confounding, providing methodological transparency for ECG-derived age estimation To support deployment in wearable devices, we benchmarked the performance of the foundation model-derived predictions across 12-lead, 6-lead, and single-lead configurations to assess their feasibility in reduced-lead settings. Finally, to evaluate the clinical utility and prognostic value of this ECG-derived age deviation, we verified our findings on the external cohort MIMIC-IV-ECG^18^, assessing the association of ECG age deviation with long-term mortality and survival outcomes. We have also developed a web application at https://bioinformatics.mdc-berlin.de/ECGage that allows users to extract biological age, age deviation, and foundation model task probabilities from provided ECG images or raw signals.

**Fig. 1.**
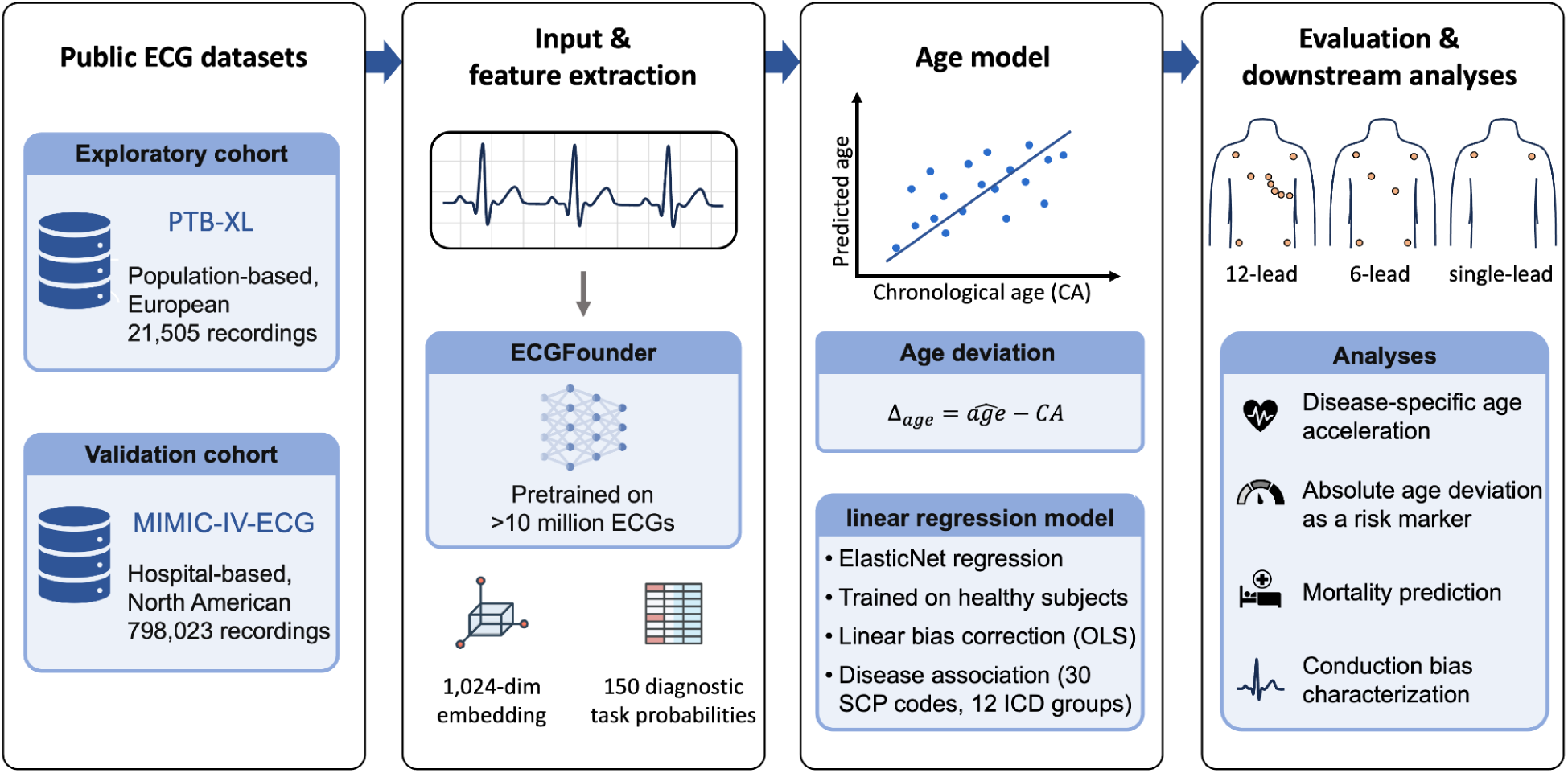
| Study workflow for ECG-derived biological age estimation. Two publicly available ECG datasets from PhysioNet. PTB-XL and MIMIC-IV-ECG were processed through ECGFounder, a foundation model pretrained on over 10 million ECGs. For each recording, 1,024-dimensional embeddings and 150 diagnostic task probabilities were extracted. An ElasticNet regression model trained exclusively on healthy subjects predicts biological age from the embeddings, with linear bias correction applied to remove age-dependent systematic error. The pipeline was independently evaluated across 12-lead, 6-lead, and single-lead configurations. Downstream analyses include disease-specific age acceleration, absolute age deviation as a universal risk marker, mortality prediction, and conduction bias characterization.

## Results

### Regression on foundation model embeddings predicts age accurately

As the basis for calculating age deviation, we established an ECG-based age prediction model using ECGFounder embeddings from healthy subjects in the PTB-XL dataset. The PTB-XL dataset comprised 21,505 recordings from 18,611 patients (11,286 male and 10,219 female recordings; 9,582 male and 9,029 female patients; mean age 59.5 years). Of these, 9,407 (43.7%) were classified as healthy, if the Standard Communication Protocol for computer-based ECG (SCP-ECG) NORM annotation probability provided with the dataset is above 50% (Figure 2a,b). A preliminary comparison of linear and non-linear ensemble regression approaches showed comparable age prediction accuracy, with linear models matching the performance of Gradient Boosting, a non-linear ensemble method (R²=0.60 vs 0.60; Supplementary Figure S1). This motivated the use of ElasticNetCV, a linear regression model that combines L1 and L2 regularization with data-adaptive hyperparameter selection.

**Fig. 2.**
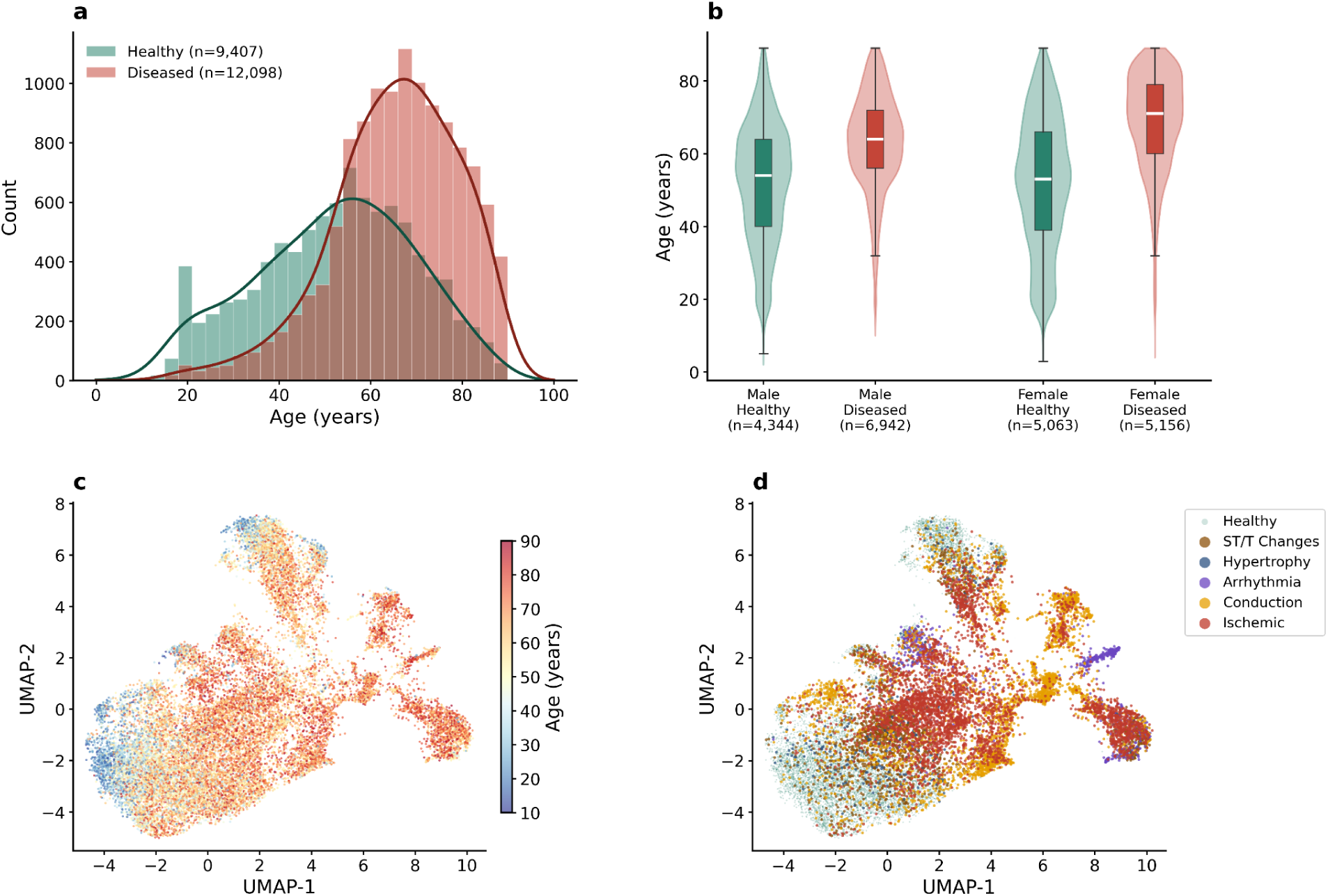
| PTB-XL cohort characteristics and ECGFounder embedding space. (a) Age distribution of healthy (n=9,407) and diseased (n=12,098) recordings with kernel density estimation overlay. Healthy recordings were defined bySCP-ECG NORM annotation probability ≥50%. Diseased subjects are shifted toward older ages (mean 65.4 vs 52.0 years). (b) Age distribution stratified by sex and health status, shown as violin plots with embedded box plots (median: white line; interquartile range: box; whiskers: 1.5× IQR). Recordings from diseased patients show an older age distribution across both sexes, with similar distributions between males and females. (c) UMAP projection of 1,024-dimensional ECGFounder embeddings (n=21,505 recordings) colored by chronological age, revealing a continuous age gradient from younger (blue, lower-left) to older (red, upper-right) patients. (d) Same UMAP projection colored by disease category based on SCP-ECG diagnostic superclasses. Conduction abnormalities and arrhythmias form distinct clusters, while ischemic conditions and ST/T changes occupy regions associated with older age in panel c, consistent with their positive age deviation.

Trained exclusively on the healthy subset with a patient-level 80:20 split (7,503 train / 1,904 test), the model achieved an R² of 0.611 and mean absolute error (MAE) of 8.31 years on the held-out test set using 12-lead embeddings (α=0.144, L1 ratio=0.50). To confirm that performance was not dependent on a single train-test split, we additionally performed five-fold patient-level cross-validation, which showed similar results and supported performance robustness (mean R²=0.624±0.012, MAE=8.25±0.13 years) (Figure 3).

**Fig. 3.**
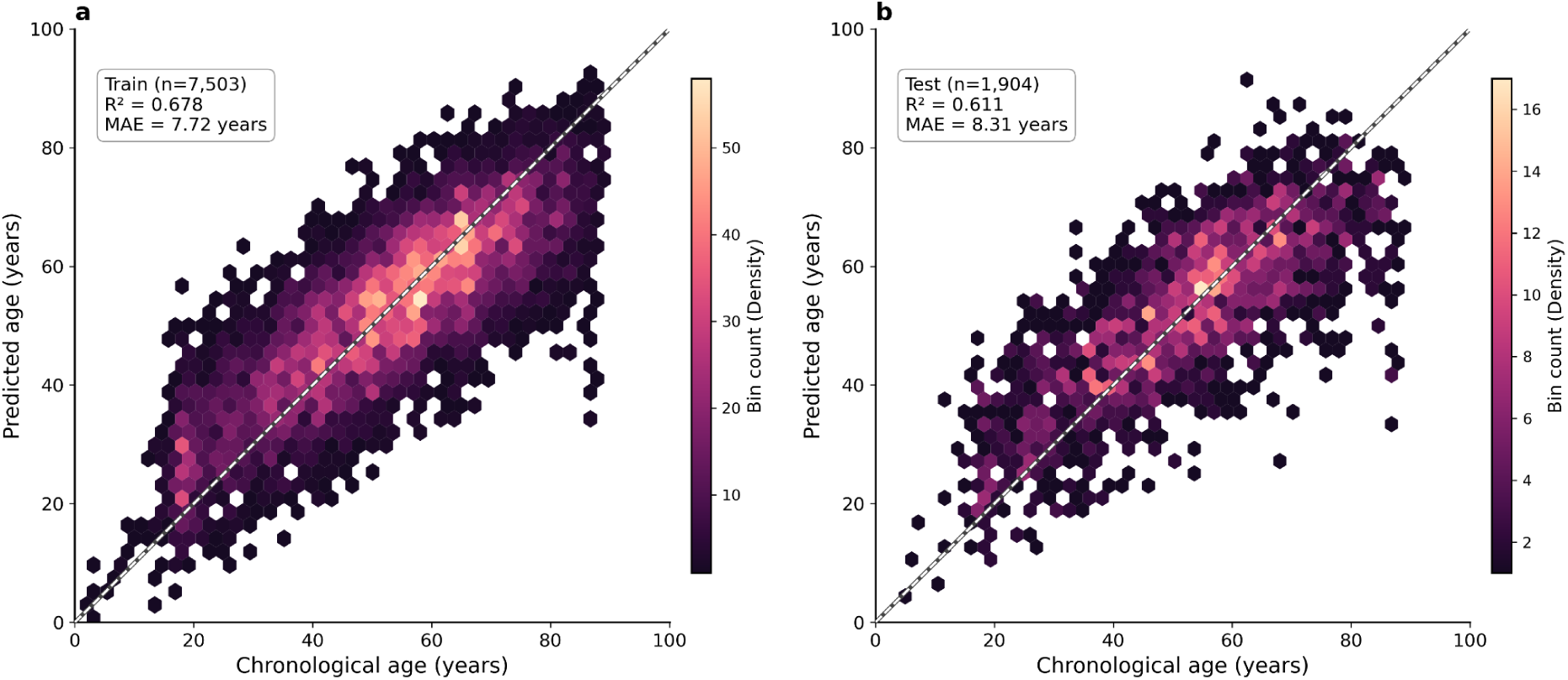
| Age prediction performance of ElasticNetCV trained on healthy PTB-XL subjects. Hexbin density plots of predicted versus chronological age for (a) training set (n=7,503; R²=0.678, MAE=7.72 years) and (b) held-out test set (n=1,904; R²=0.611, MAE=8.31 years). Color intensity represents point density. The dashed line indicates perfect prediction (y=x). The model was trained exclusively on healthy subjects with patient-level splitting to prevent data leakage. The modest performance decrease from training to test confirms generalization without overfitting. ElasticNetCV was selected over Ridge, Lasso, Random Forest, and Gradient Boosting based on comparable accuracy with greater interpretability (Supplementary Figure S1).

To account for a systematic bias in the age prediction toward the mean, a linear ordinary least squares (OLS) correction was applied to the predicted ages. Following correction, the relationship between predicted and chronological age was approximately linear (Pearson r=0.783, Spearman r=0.782), and the residual slope on the test set was near zero (−0.038), confirming effective reduction of the systematic age-dependent bias. A comparison of linear, quadratic, and spline correction models in MIMIC-IV-ECG confirmed negligible differences (R²: 0.563, 0.566, 0.566; residual slopes: 0.0001, 0.0000, 0.0000; Supplementary Figure S5), indicating that linear OLS correction is sufficient and that age deviation values could be interpreted without further calibration.

Consistent with the regression results, Uniform Manifold Approximation and Projection (UMAP) visualization of the 1,024-dimensional embedding space revealed a clear age gradient, with younger patients clustering separately from older patients (Figure 2c). Additional coloring by disease category showed distinct clustering of conduction abnormalities and arrhythmias, while ischemic conditions and ST-segment/T-wave (ST/T) changes occupied regions associated with older age (Figure 2d). Together, these findings suggest that ECGFounder embeddings encode aging-related information, supporting their use for subsequent age deviation analyses.

### Disease-associated age acceleration

After establishing the age prediction model, we next examined disease-associated age deviation. Because conduction abnormalities can substantially alter ECG morphology, we analyzed non-conduction diagnoses separately. Among these diagnoses, diseased subjects exhibited significantly higher age acceleration than healthy controls, with an overall delta of +3.33 years (95% bootstrap CI: 2.87–3.79, Cohen’s d=0.332). Here, age acceleration refers to positive age deviation, where ECG-derived age exceeds CA. This pattern was consistent across 30 individual SCP codes tested (n≥100 each)after false discovery rate (FDR) correction. The strongest effects were observed for digitalis effect (DIG: Δ=+6.32y, d=0.673), anterolateral ischemia (ISCAL: Δ=+6.15y, d=0.638), and long QT interval (LNGQT: Δ=+6.09y, d=0.696) (Figure 4a). At the disease category level, ST/T changes showed the largest acceleration (Δ=+4.81y, d=0.517), followed by ischemic conditions (Δ=+4.31y, d=0.444) and hypertrophy (Δ=+4.11y, d=0.408) (Figure 4d).

**Fig. 4.**
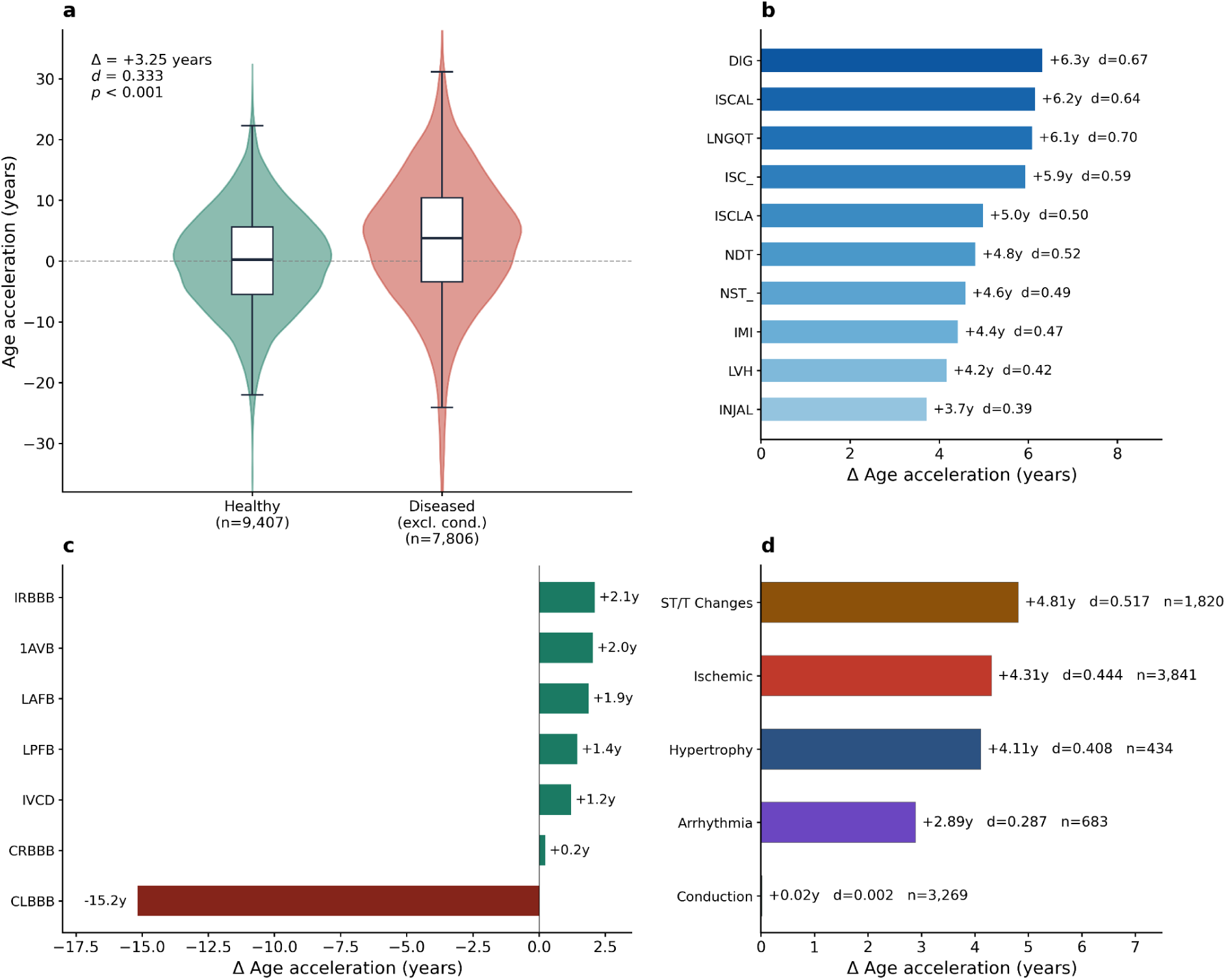
| Disease-specific age acceleration in PTB-XL. (a) Age acceleration comparison between healthy individuals (n=9,407) and diseased subjects (n=7,806, excluding conduction abnormalities). Diseased subjects exhibit a significantly higher mean age acceleration compared to healthy controls (Δ = +3.25 years, Cohen’s d = 0.333, p < 0.001).(b) Top 10 non-conduction diagnoses ranked by mean age acceleration relative to healthy controls (all FDR-corrected p<0.001). Digitalis effect (DIG: Δ=+6.3y, d=0.67), anterolateral ischemia (ISCAL: Δ=+6.2y, d=0.64), and long QT interval (LNGQT: Δ=+6.1y, d=0.70) showed the strongest effects. Bar color intensity reflects effect magnitude. (c) Conduction abnormality subtypes analyzed separately. Most conduction subtypes showed mild positive acceleration consistent with age-related conduction slowing, while complete left bundle branch block (CLBBB) exhibited a striking negative deviation of −15.2 years, indicating that the ECGFounder embedding space encodes CLBBB morphology as a younger ECG pattern. (d) Disease category summary across five SCP-ECG superclasses. ST/T changes (Δ=+4.81y, d=0.517) and ischemic conditions (Δ=+4.31y, d=0.444) showed the largest category-level effects. Conduction as a category showed near-zero acceleration (Δ=+0.02y, d=0.002) due to the opposing effects of CLBBB and other conduction subtypes. Sample sizes (n) and Cohen’s d effect sizes are annotated for each category.

Notably, conduction abnormalities as a category showed negligible age acceleration (Δ=+0.02y, d=0.002, p=0.95). This near-zero effect was driven by opposing effects of individual subtypes. Complete left bundle branch block (CLBBB) showed negative age deviation (Δ=−15.18y, d=−1.515) while other conduction subtypes showed mild positive deviation, including incomplete right bundle branch block (IRBBB: Δ=+2.11y) and first-degree atrioventricular block(1AVB: Δ=+2.03y) (Figure 4b). This conduction-specific pattern, in which a single diagnostic subtype reverses the direction of age acceleration for an entire disease category, represents a previously uncharacterized confound in foundation model embeddings, with implications examined in the cross-cohort comparison below.

### Reduced-lead configurations retain disease discrimination

To assess the feasibility of ECG-derived age deviation in wearable devices, we next compared the standard 12-lead configuration with 6-lead and single-lead configurations. As expected, reduced-lead configurations showed a decline in age prediction accuracy. Six-lead embeddings achieved R²=0.514 and MAE=9.29 years, while single-lead embeddings achieved R²=0.494 and MAE=9.57 years. However, disease-versus-healthy discrimination was largely preserved across lead configurations: 6-lead performance (d=0.337) slightly exceeded 12-lead performance (d=0.332), while single-lead performance showed only modest degradation (d=0.304). Additionally, the disease-versus-healthy delta was larger for reduced-lead configurations (6-lead: +3.51y; 1-lead: +3.44y) compared to 12-lead configuration (+3.33y). Because the reduced 6-lead configuration does not include the precordial chest leads available in the standard 12-lead ECG, this pattern suggests that the precordial leads contribute additional age-predictive information but may also introduce morphological confounds that dilute the disease discrimination signal (Figure 5a–c).

**Fig. 5.**
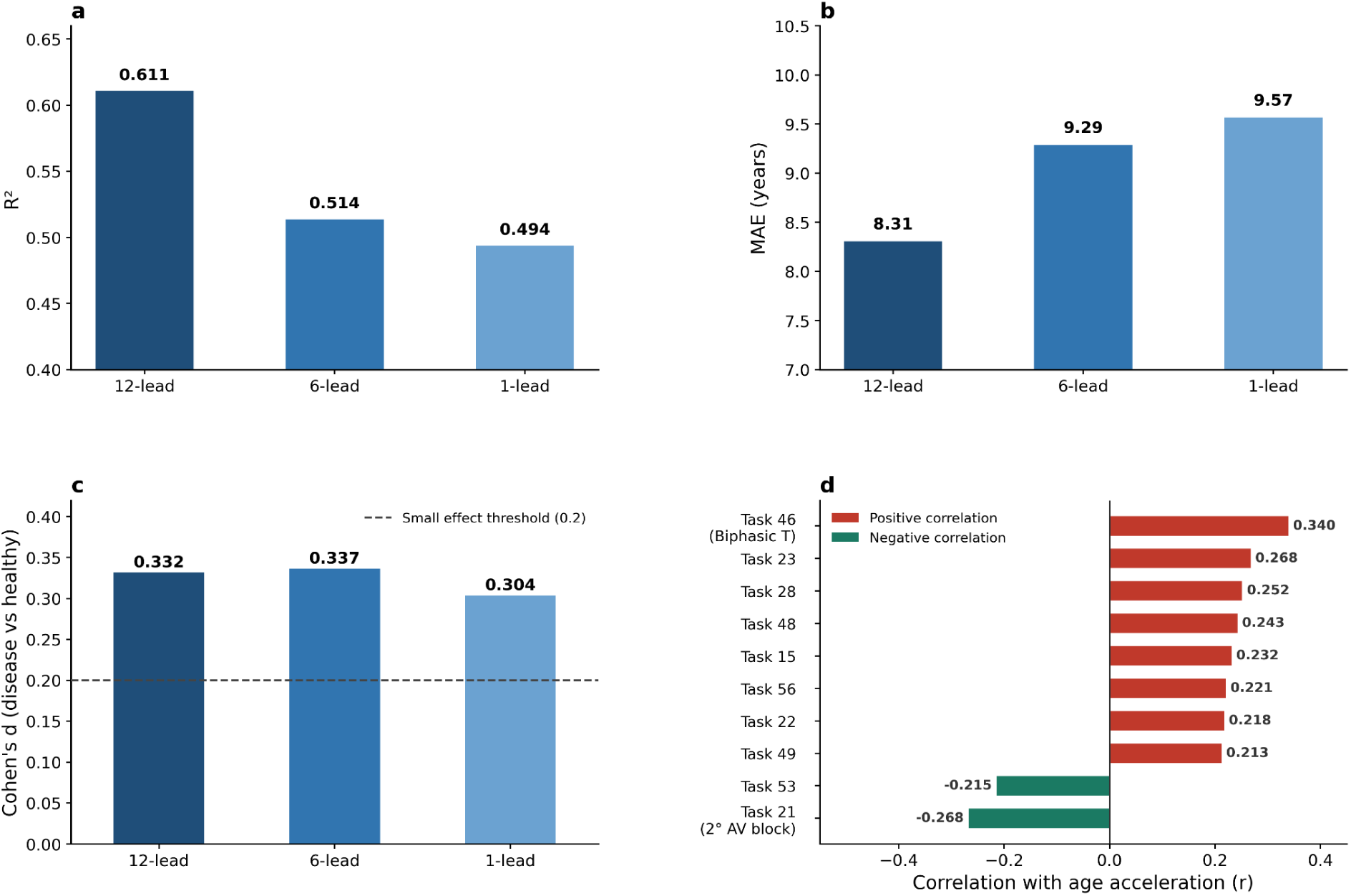
| Lead ablation and task probability correlations in PTB-XL. (a) Age prediction accuracy (R²) across three lead configurations: 12-lead (0.611), 6-lead (0.514), and single-lead (0.494). For 6-lead, each limb lead was independently processed through the 1-channel ECGFounder model and embeddings were averaged. (b) Mean absolute error (MAE) increases with fewer leads, from 8.31 years (12-lead) to 9.57 years (single-lead). (c) Disease-versus-healthy discrimination (Cohen’s d) was largely preserved across configurations: 6-lead (d=0.337) slightly exceeded 12-lead (d=0.332), while single-lead (d=0.304) showed only modest degradation. The dashed line indicates the conventional small effect threshold (d=0.2). This suggests that precordial leads contribute age-predictive information but may introduce morphological confounds that dilute disease discrimination. (d) Top 10 ECGFounder task probabilities correlated with age acceleration (FDR-corrected). Biphasic T waves (Task 46, r=0.340) showed the strongest positive correlation, while second-degree AV block Type I (Task 21, r=−0.268) showed the strongest negative correlation, consistent with the conduction bias pattern observed in Figure 4.

To explore ECG features associated with age acceleration, we correlated age acceleration with the 150 diagnostic task probabilities generated by ECGFounder. Among the 150 ECGFounder task probability outputs, biphasic T waves (Task 46) showed the strongest positive correlation with age acceleration (r=0.340), whereas second degree AV block Type I (Task 21) showed the strongest negative correlation (r=−0.268), consistent with the observed conduction bias pattern (Figure 5d). Sex-stratified analysis revealed that females consistently showed higher R² than males across all configurations (12-lead: 0.643 vs 0.564; 6-lead: 0.532 vs 0.488; single-lead: 0.526 vs 0.448), suggesting a more consistent aging signal in female ECG embeddings (Supplementary Figure S4).

### External validation in MIMIC-IV-ECG

After establishing the age prediction pipeline and characterizing disease-specific acceleration patterns in the PTB-XL dataset, a population-based European cohort, we next evaluated whether these findings generalize to an independent clinical setting with distinct patient demographics and disease burden. The MIMIC-IV-ECG dataset is a hospital-based North American cohort nearly 40-fold larger than PTB-XL and additionally provides linked mortality outcomes, enabling direct assessment of prognostic value.

In the healthy subset of MIMIC-IV-ECG (127,742 recordings from 60,181 patients), ElasticNetCV achieved R²=0.544 and MAE=8.81 years, with near-identical train and test performance (train R²=0.558, test R²=0.544) indicating no overfitting (Figure 6a). The lower R² compared to PTB-XL likely reflects the greater heterogeneity of a hospital-based cohort and inherent age uncertainty from the MIMIC-IV de-identification procedure. Lead ablation confirmed the PTB-XL pattern, with lower accuracy in reduced-lead configurations: 6-lead achieved R²=0.499 (MAE=9.25) and single-lead achieved R²=0.449 (MAE=9.69) (Figure 6b).

**Fig. 6.**
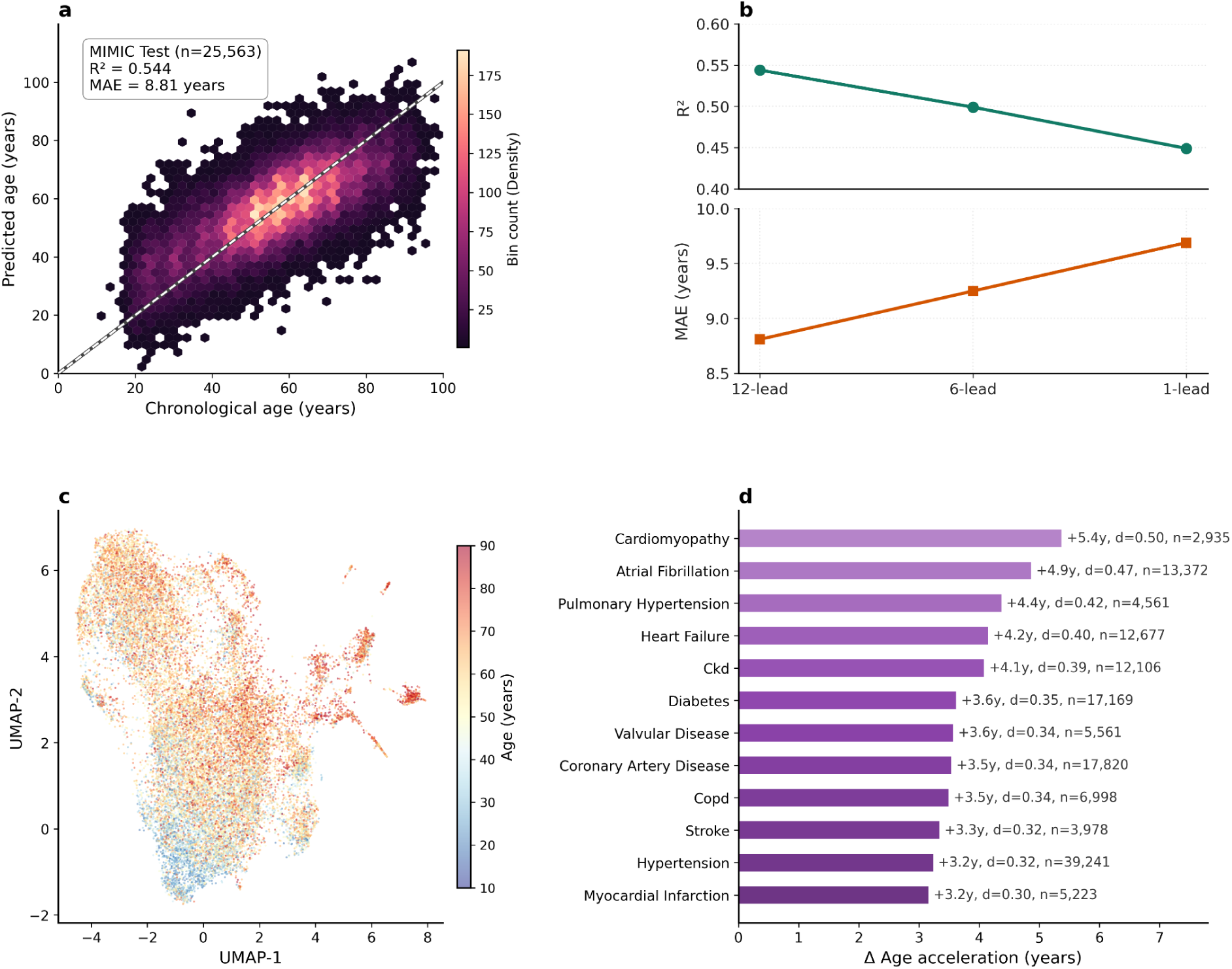
| External validation in MIMIC-IV-ECG. (a) Hexbin density plot of predicted versus CA on the MIMIC-IV-ECG test set (n=25,563; R²=0.544, MAE=8.81 years). The lower R² compared to PTB-XL (0.611) likely reflects the greater heterogeneity of a hospital-based cohort and inherent age uncertainty from the MIMIC-IV de-identification procedure. The dashed line indicates a perfect prediction. (b) Lead ablation in MIMIC-IV-ECG is consistent with the PTB-XL pattern: R² decreased from 0.544 (12-lead) to 0.499 (6-lead) and 0.449 (single-lead), with corresponding MAE increases from 8.81 to 9.69 years. (c) UMAP projection of MIMIC-IV-ECG ECGFounder embeddings (n=25,000 random subset) colored by chronological age, showing a clear age gradient consistent with the PTB-XL embedding structure shown in Figure 2c. (d) All 12 ICD-10-based disease groups showed significant positive age deviation after FDR correction. Cardiomyopathy exhibited the highest positive deviation (Δ=+5.4y, d=0.50, n=2,935), followed by atrial fibrillation (Δ=+4.9y, d=0.47, n=13,372) and pulmonary hypertension (Δ=+4.4y, d=0.42, n=4,561). The disease hierarchy was broadly consistent with PTB-XL, with structural cardiac conditions and electrical disorders showing the largest effects in both cohorts.

All 12 International Classification of Diseases, 10th Revision (ICD-10)-based disease groups showed significant positive age deviation after FDR correction (Figure 6d). ICD-10-based disease groups showed significant positive age deviation after FDR correction (Figure 6d). Cardiomyopathy exhibited the highest positive deviation (n=2,935 patients; Δ=+5.37y, d=0.497), followed by atrial fibrillation (n=13,372; Δ=+4.87y, d=0.470), pulmonary hypertension (n=4,561; Δ=+4.37y, d=0.417), and heart failure (n=12,677; Δ=+4.15y, d=0.396). Systemic conditions including chronic kidney disease (n=12,106; Δ=+4.09y, d=0.394) and diabetes (n=17,169; Δ=+3.62y, d=0.354) also showed significant positive deviation. Consistent with the PTB-XL lead ablation findings, reduced-lead configurations in MIMIC maintained overall age prediction and mortality prediction. However, disease-specific analysis revealed that single-lead recordings showed paradoxical negative deviation for several conditions including hypertension (Δ=−0.95y) and diabetes (Δ=−0.68y). This suggests that while single-lead ECGs are sufficient for overall risk assessment, disease-specific interpretation should be restricted to 6-lead or 12-lead recordings.

UMAP visualization of the MIMIC-IV-ECG embedding space further supports that age- and disease-related information is encoded in the foundation model representations of ECGFounder, with clear age gradients and distinct disease-category clusters (Figure 6c).

### Age deviation predicts mortality in younger patients

To determine whether ECG-derived age deviation carries prognostic information, we assessed its association with all-cause mortality in MIMIC-IV-ECG. Cox proportional hazards regression on 160,493 MIMIC-IV-ECG patients, including 26,902 deaths (16.8%), showed that age deviation was associated with all-cause mortality after adjustment for CA and sex. Each 1-year increase in age deviation was associated with a 0.5% higher mortality risk (HR=1.005, 95% CI: 1.004–1.006, p=1.88×10⁻¹⁷).

Age deviation was most informative in patients younger than 65 years (n=100,085; HR=1.014, p=6.21×10⁻³⁸), with weaker effects in patients aged 65–75 years (HR=1.005, p=3.70×10⁻⁵) and no significant association in patients older than 75 years (HR=0.999, p=0.193), according to Table 1. The association was also stronger in males (HR=1.008, p=8.68×10⁻²²) than in females (HR=1.002, p=0.019).

**Table 1.**
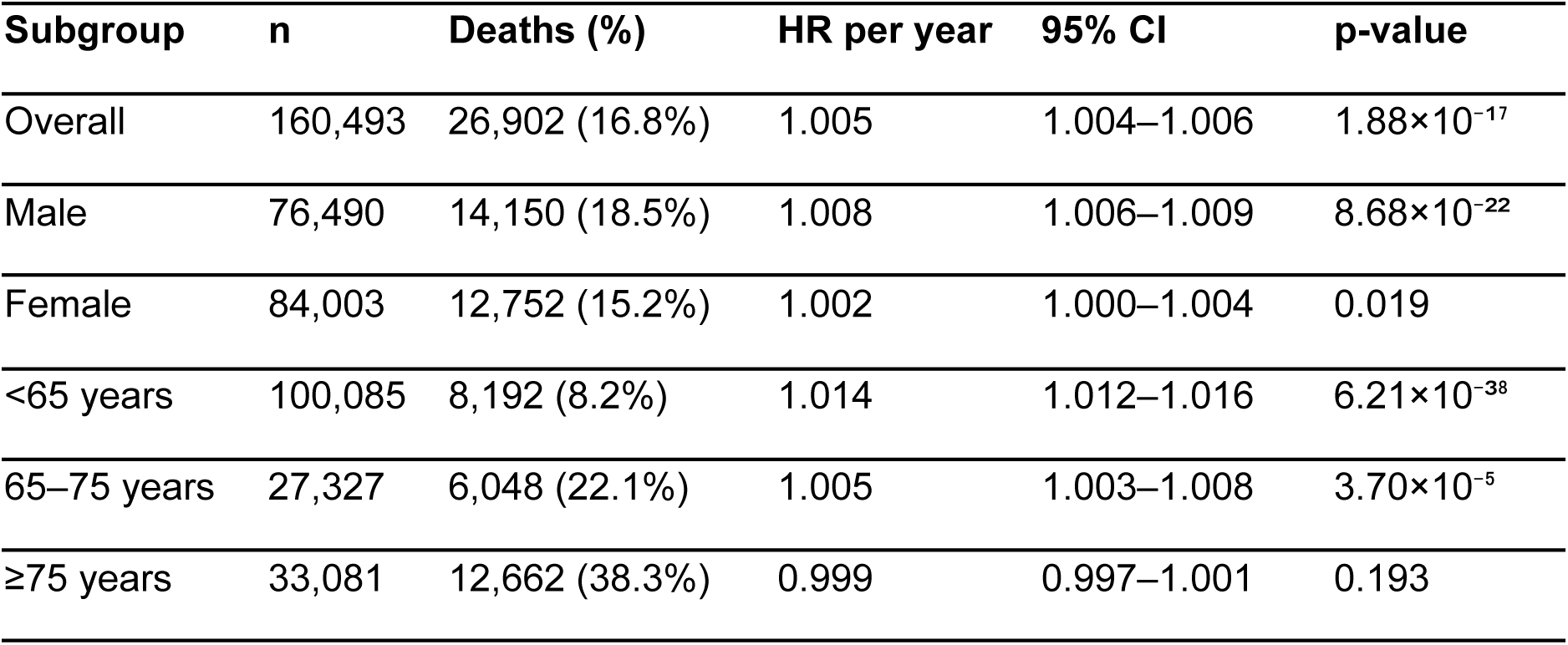
Association between ECG-derived age deviation and all-cause mortality in MIMIC-IV-ECG, stratified by sex and age group. Cox proportional hazards regression was adjusted for CA and sex. Hazard ratios (HR) are reported per 1-year increase in age acceleration with 95% confidence intervals. The strongest association was observed in patients under 65 years (HR=1.014), while no significant association was found in patients over 75 (HR=0.999, p=0.193), suggesting that age deviation is most clinically informative as a screening tool in younger populations where CA alone is a weaker predictor of cardiac risk.

| Subgroup | n | Deaths (%) | HR per year | 95% CI | p-value |
| --- | --- | --- | --- | --- | --- |
| Overall | 160,493 | 26,902 (16.8%) | 1.005 | 1.004–1.006 | $1.88 \times 10^{-17}$ |
| Male | 76,490 | 14,150 (18.5%) | 1.008 | 1.006–1.009 | $8.68 \times 10^{-22}$ |
| Female | 84,003 | 12,752 (15.2%) | 1.002 | 1.000–1.004 | 0.019 |
| <65 years | 100,085 | 8,192 (8.2%) | 1.014 | 1.012–1.016 | $6.21 \times 10^{-38}$ |
| 65–75 years | 27,327 | 6,048 (22.1%) | 1.005 | 1.003–1.008 | $3.70 \times 10^{-5}$ |
| ≥75 years | 33,081 | 12,662 (38.3%) | 0.999 | 0.997–1.001 | 0.193 |

Landmark analyses were performed to assess whether the mortality association was driven by acute peri-recording events. The hazard ratio increased progressively from 1.005 at baseline to 1.007 at 30 days, 1.009 at 90 days, and 1.010 at 180 days after ECG acquisition (Supplementary Table S6), suggesting that the association was not explained by deaths occurring shortly after ECG recording.

Adding age deviation to a baseline model including CA and sex produced a small increase in C-index (+0.0004). Age deviation alone achieved a C-index of 0.510, compared with 0.743 for CA alone, 0.745 for age plus sex, and 0.746 for the full model including age deviation (95% CI: 0.744–0.748). This small increase in the C-index is consistent with the known insensitivity of C-index to added predictors, even when those predictors are statistically significant and clinically relevant, particularly when the baseline model already includes a dominant predictor such as CA^19^. Age deviation, by design, captures whether an individual’s ECG-derived age is higher or lower than expected for their CA, identifying individuals who appear to age faster or slower than their peers. Its prognostic value was therefore more evident within age strata, particularly among patients younger than 65, in whom each 1-year increase in age deviation was associated with higher mortality risk (HR=1.014; p=6.21×10⁻³⁸). Together, the significant association in the overall Cox model (p=1.88×10⁻¹⁷) and progressive increase in hazard ratios in landmark analyses confirm that age deviation carries independent prognostic information beyond CA alone.

Schoenfeld residual tests indicated potential violation of the proportional hazards assumption for CA (p=3.21×10⁻²⁵) and age deviation (p=6.43×10⁻³⁸), suggesting that their associations with mortality may vary over follow-up time. Given the large sample size, these tests may detect even small deviations from proportionality. Therefore, the landmark analyses described above were used to assess the robustness of the mortality association over time.

### Cross-cohort conduction analysis supports absolute age deviation as a universal risk marker

The consistency of findings across PTB-XL (population-based, European, 1989–1996) and MIMIC-IV-ECG (hospital-based, North American, contemporary) strengthens the generalizability of ECG-derived biological age as a biomarker. Both cohorts showed significant disease-associated age acceleration across multiple diagnostic categories, with structural and ischemic heart diseases producing the largest effects.

However, conduction abnormalities exhibited a striking cross-cohort divergence. In PTB-XL, conduction abnormalities showed near-zero acceleration (Δ=+0.02y) driven by the dominant negative effect of CLBBB (Δ=−15.18y). This pattern suggests a morphology-related confound in the embedding space, supported by the three-configuration sensitivity analysis (Supplementary Figure S2). High-error analysis further identified CLBBB (7.05-fold enrichment) and pacemaker rhythm (4.34-fold) as the dominant sources of prediction error in PTB-XL (Supplementary Figure S3). In contrast, full-population conduction analysis in MIMIC-IV-ECG (127,237 conduction vs. 543,413 non-conduction recordings) revealed the opposite pattern, with conduction patients showing strong positive acceleration (Δ=+5.93y, d=0.557). This divergence reflects two complementary phenomena. In PTB-XL, conduction findings are often isolated morphological variants where CLBBB dominates the category average. In MIMIC-IV-ECG, conduction abnormalities co-occur with significant comorbidity burden. Additionally, MIMIC-IV-ECG identifies conduction via report keywords that do not distinguish CLBBB from other bundle branch blocks, preventing isolation of the morphological confound using the more granular SCP-ECG coding in PTB-XL.

This cross-cohort comparison highlights a limitation of signed age deviation as a biomarker: large deviations in opposite directions may arise from different mechanisms, including morphology-related confounding reflected by negative deviation and comorbidity-driven aging reflected by positive deviation. Because both indicate that ECG-derived age differs substantially from chronological age, we therefore propose absolute age deviation (|predicted age − chronological age|) as a more robust universal risk marker, as it flags both directions of abnormality under a single metric.

Additional analyses in MIMIC-IV-ECG revealed systematic variation in prediction accuracy across ethnicity groups. Asian patients showed the largest deviation (Δ=−2.84y, d=−0.266, FDR-significant), whereas Hispanic patients showed positive deviation (Δ=+1.53y). Black (n=19,442) and Other (n=5,031) groups were statistically underpowered, highlighting the need for ethnicity-aware calibration in clinical deployment. Temporal stability analysis among 103,880 patients with multiple ECGs showed moderate within-patient consistency (ICC=0.499, within-patient SD=6.40 years), with minimal systematic drift over time (median longitudinal slope=−0.14 years per year).

## Discussion

Our findings demonstrate that the foundation model embeddings encode sufficient biological information to predict ECG-derived age, and that the resulting age deviation serves as a robust, cross-cohort biomarker for cardiovascular diseases. Using ECGFounder embeddings together with a linear model can effectively capture clinically meaningful age estimates (R²=0.611, MAE=8.31 years in PTB-XL; R²=0.544, MAE=8.81 years in MIMIC-IV-ECG) without requiring complex downstream architectures, yielding results comparable to studies employing end-to-end deep learning architectures trained on millions of ECGs ^20,21^. This is coherent with the broader paradigm shift toward foundation models in medical AI, where pretrained representations increasingly replace the need for task-specific model development ^22^.

The consistent positive age deviation observed across disease categories in both cohorts supports the interpretation that ECG-derived BA reflects genuine pathophysiological burden rather than statistical artifact. The disease hierarchy was biologically coherent: conditions involving structural remodeling (cardiomyopathy, hypertrophy), electrical dysfunction (atrial fibrillation), and ischemic injury showed the largest effects, consistent with the known impact of these processes on cardiac electrophysiology and the hallmarks of cardiovascular aging recently outlined in comprehensive reviews^22,23^. The concept of accelerated cardiac aging as a consequence of repeated pathological stress has been proposed in congenital heart disease^24^. Our findings extend this framework to acquired cardiovascular conditions in adult populations, providing a non-invasive, ECG-based quantification of this phenomenon consistent with the cardiac aging biomarker framework proposed by the Aging Biomarker Consortium^25^.

Notably, systemic conditions including chronic kidney disease and diabetes also showed significant age acceleration in MIMIC-IV-ECG, supporting the emerging concept that extracardiac diseases contribute to measurable cardiac aging through shared pathways of inflammation, metabolic stress, and vascular dysfunction^26^. Recent studies have demonstrated that deep learning models can detect chronic kidney disease directly from ECG signals^27,28^, and that ECG-based algorithms can identify electrolyte disturbances associated with renal dysfunction^29^. These findings suggest that the ECGFounder embedding space captures subtle electrophysiological signatures of systemic diseases that can manifest as apparent cardiac aging. Similarly, the significant diabetes-associated acceleration observed in our study parallels recent evidence linking accelerated biological aging to incident cardiovascular disease in diabetic populations^30,31^ (Figure 6d).

The similar age prediction performance and disease-acceleration patterns observed in both the exploratory cohort PTB-XL (population-based, European, 1989–1996) and the external validation cohort MIMIC-IV-ECG (hospital-based, North American, contemporary) support the generalizability of these findings. Despite substantial differences in patient demographics, disease prevalence, recording technology, and diagnostic labeling frameworks, the overall pattern of disease-associated cardiac aging was consistent. This cross-cohort robustness positions ECG-derived age deviation alongside other biological aging measures, including epigenetic clocks, phenotypic age, and clinical biomarker-derived indices as a complementary modality for cardiovascular risk assessment^32^. Notably, our ECG-based approach offers practical advantages over blood-based aging biomarkers, as it requires no laboratory processing, provides results in seconds, and can be obtained from existing clinical recordings without additional costs.

Our analysis of conduction abnormalities reveals an important limitation for the interpretation of embedding-based age biomarkers. In PTB-XL, CLBBB produced a negative age deviation of −15.18 years, likely reflecting the model’s association of CLBBB-related ECG morphology with patterns more commonly observed in younger individuals. Our three-configuration sensitivity analysis confirmed that these confounders are intrinsic to the embedding space and cannot be corrected by adding task probabilities or disease labels (Supplementary Figure S2). In contrast, conduction abnormalities in MIMIC-IV-ECG showed strong positive acceleration (+5.93 years), reflecting the comorbidity burden in a hospital-based cohort. This cross-cohort divergence highlights the need for morphology-aware interpretation and motivates our proposal of absolute age deviation (|ŷ − f(y)|, where f(y) is the OLS-corrected expected age) as a more robust metric that captures abnormality in both directions. To our knowledge, this is the first systematic characterization of conduction-induced confounding in foundation model embeddings applied to age prediction, and it carries implications beyond ECG analysis for any embedding-based biomarker derived from signals with morphologically distinct subtypes.

The preservation of disease discrimination and mortality prediction across reduced-lead configurations has important implications for scalable cardiovascular screening. Six-lead recordings, compatible with consumer devices such as KardiaMobile ^33^ (example studies: ^34,35^) retained disease discrimination comparable to the full 12-lead configuration (Cohen’s d=0.337 versus 0.332), while single-lead recordings still achieved significant mortality prediction (HR=1.003, p=1.01×10⁻³⁰). These findings are consistent with the growing evidence that wearable ECG devices provide clinically actionable cardiac information^14^ and with concurrent work by Dhingra et al.^36^ demonstrating that single-lead ECG-age from Apple Watch and KardiaMobile devices predicts major adverse cardiovascular events across multiple cohorts. Our foundation model approach complements their end-to-end architecture by offering a systematic comparison across lead configurations and enabling direct analysis of the embedding space, which revealed the conduction bias characterization described above. However, our finding that single-lead configuration produced paradoxical negative deviation for hypertension and diabetes in MIMIC-IV-ECG indicates that disease-specific interpretation should be limited to 6-lead or 12-lead recordings.

The mortality analysis revealed that ECG-derived age deviation is most prognostic in younger patients. The hazard ratio of 1.014 per year in patients under 65 (p=6.21×10⁻³⁸) suggests that this biomarker is particularly valuable for identifying premature cardiovascular aging before CA dominates risk prediction. In contrast, the non-significant association in patients over 75 likely reflects the overwhelming influence of other risk factors and comorbidities in elderly populations. This age-dependent prognostic pattern is consistent with phenotypic age acceleration studies showing stronger mortality associations in younger cohorts^3,37^, and with ECG-based mortality prediction models reporting the strongest discrimination in middle-aged populations^21^. The progressive strengthening of the hazard ratio in landmark analysis (1.005 at baseline to 1.010 at 180 days) confirms that the mortality association reflects long-term prognostic value rather than acute illness severity.

While our findings provide robust evidence for the utility of age deviation as a risk biomarker, several methodological limitations should be considered. A primary consideration is the retrospective, observational nature of both cohorts, which precludes direct causal inference. However, the consistency of the associations across distinct populations offers a strong foundation for future mechanistic studies. Similarly, data characteristics inherent to large clinical registries introduce specific nuances. For instance, the MIMIC-IV de-identification procedure employs date-shifting, which introduces age uncertainty. While this potentially dampens our observed prediction accuracy and effect sizes, it also suggests that our reported boundaries represent conservative estimates of a resilient underlying signal.

Furthermore, the operational definitions and diagnostic frameworks varied between datasets, utilizing expert-assigned SCP codes in PTB-XL versus billing-derived ICD-10 codes and report keywords in MIMIC-IV. Although this algorithmic diversity introduces classification noise and limitations in fully capturing “healthy” reference states, the resilience of the age deviation signal across such disparate clinical documentation styles underscores its generalizability. Finally, our sub-analyses highlight critical directions for future clinical deployment. While ethnicity analyses revealed systematic variations, which is most notable in Asian patients (Δ=−2.84 years), other cohorts remained statistically underpowered, emphasizing the urgent need for larger, more diverse datasets to ensure algorithmic equity. Moreover, while absolute age deviation emerged as a compelling universal marker, validating its prospective prognostic value against hard mortality outcomes and investigating the incremental benefit of serial measurements (ICC=0.499) remain essential next steps to increase its clinical utility.

## Methods

### Study Design

We conducted a two-cohort observational study to investigate ECG-derived biological age using embeddings from a pretrained foundation model. The primary development cohort was PTB-XL, a population-based ECG dataset used for model development, disease-specific age acceleration analysis, conduction bias investigation, and lead configuration comparison. External validation was performed on MIMIC-IV-ECG, a hospital-based clinical dataset, where we additionally assessed the prognostic value of ECG-derived age acceleration for all-cause mortality.

### Datasets

#### PTB-XL

PTB-XL is a publicly available dataset comprising 21,837 twelve-lead clinical ECG recordings from 18,885 patients of 10-second length, collected at the University Hospital of Freiburg between 1989 and 1996 [16]. Each recording is annotated by up to two cardiologists with diagnostic labels from 71 SCP-ECG statements, along with patient demographics (age, sex) and a unique patient identifier enabling patient-level analyses. After ECGFounder embedding extraction (21,799 successful), removal of one record with missing embedding values, and exclusion of 293 recordings with age exceeding 100 years, 21,505 recordings from 18,611 unique patients remained for analysis. Of these, 9,407 (43.7%) were classified as healthy and 12,098 (56.3%) as diseased.

Healthy subjects were defined as those with a NORM (normal ECG) probability ≥50% in the SCP-ECG annotations, consistent with the dataset’s validated diagnostic labeling scheme. Disease categories were constructed from SCP codes grouped into five clinically meaningful superclasses: ischemic (ISCAL, ISCLA, ISCAS, ISCIN, ISCAN, ISC_, IMI, AMI, LMI, PMI), hypertrophy (LVH, RVH, LAO, RAO), conduction abnormalities (CLBBB, CRBBB, IRBBB, ILBBB, LAFB, LPFB, 1AVB, 2AVB, 3AVB, IVCD), arrhythmia (AFIB, AFLT, PACE, PVC, PAC, SVTAC, STACH, SBRAD), and ST/T changes (NST_, NDT, DIG, LNGQT, APTS). Individual SCP codes with at least 100 occurrences were additionally analyzed at the granular level.

#### MIMIC-IV-ECG

MIMIC-IV-ECG is a large-scale clinical ECG dataset containing 800,000 twelve-lead recordings from approximately 160,000 patients in the Beth Israel Deaconess Medical Center intensive care system [17]. Clinical metadata; including age, sex, ethnicity, date of death, and free-text machine-generated ECG reports, was linked from the MIMIC-IV clinical database by merging ECG records with the patients, admissions, and diagnoses tables via subject_id. ICD-10 diagnosis codes were extracted from the diagnoses_icd table.

Ages were computed as anchor_age + (ecg_year − anchor_year), where ecg_year is the year of ECG recording extracted from the ecg_time field, and clipped to the range [0, 100]. Ages carry inherent uncertainty due to the MIMIC-IV de-identification procedure, in which dates are shifted by a random offset per patient. Date of death was extracted from the core/patients table. After merging embeddings with clinical metadata via an inner join on study_id, 798,023 recordings remained (>99.9% retention).

Healthy subjects in MIMIC-IV-ECG were identified using a keyword-based approach applied to the machine-generated ECG report text. Reports containing any of 22 abnormality-indicating keywords including atrial fibrillation, bundle branch block, ST elevation/depression, hypertrophy, ischemia, pacemaker, and others were classified as abnormal. Reports free of all such keywords were classified as healthy (*n=127,742; 16.0%*). Conduction abnormalities were identified by a separate set of conduction-specific keywords in the report text (e.g., “bundle branch block”, “fascicular block”, “AV block”), yielding 127,462 conduction-flagged recordings (15.9%).

Diseases were identified by matching ICD-10 codes against curated prefix lists for 12 cardiovascular and systemic conditions: hypertension (I10–I15), atrial fibrillation (I48), heart failure (I50), coronary artery disease (I20–I25), myocardial infarction (I21–I22), diabetes mellitus (E10–E14), cardiomyopathy (I42–I43), valvular heart disease (I34–I38), cerebrovascular disease (I60–I64), chronic kidney disease (N18–N19), and chronic obstructive pulmonary disease (J44) and and pulmonary hypertension (I27). Both emergency department (icd_code10) and hospital billing (icd_code10_hosp) ICD fields were searched, as diagnosis codes were stored as numpy arrays requiring element-wise prefix matching.

### ECG Embeddings

All ECG recordings were processed through ECGFounder, a one-dimensional convolutional neural network (Net1D architecture) pretrained on over 10 million ECGs spanning 150 diagnostic categories ^22^. For each recording, we extracted two sets of features: a 1,024-dimensional embedding vector from the penultimate fully connected layer, representing a learned high-dimensional representation of the ECG signal, and 150 task probability outputs from the softmax layer, corresponding to the model’s diagnostic predictions across its training categories.

Inference was performed with batch normalization and dropout disabled (use_bn=False, use_do=False), matching the official ECGFounder evaluation protocol as specified in the repository’s ptbxl_eval.py script. This distinction is critical: although the Net1D class defaults to use_bn=True and use_do=True, the pretrained weights were saved with these layers active during training but bypassed during inference. Using the incorrect setting would alter the forward pass and produce inconsistent embeddings.

Embeddings were extracted for three lead configurations. For 12-lead, all channels were processed through the 12-channel ECGFounder model. For single-lead, only Lead I was processed through the 1-channel model. For 6-lead, each of the six limb leads (I, II, III, aVR, aVL, aVF) was independently processed through the 1-channel model, and the resulting embeddings and task probabilities were averaged. This approach was adopted because ECGFounder provides only 12-channel and 1-channel architectures; averaging per-lead representations avoids introducing zero-padded channels outside the model’s training distribution. We note that only two of six limb leads are electrically independent (Einthoven’s law). Each configuration produces a 1,024-dimensional embedding enabling direct comparison across lead setups.

### Age Prediction Model

#### Feature Construction

For PTB-XL, the feature matrix consisted of 1,025 dimensions: 1,024 embedding features plus sex (binary: 0=male, 1=female). For MIMIC-IV-ECG, features consisted of 1,024 embedding dimensions only. Feature ablation in PTB-XL demonstrated that adding sex as a covariate yielded negligible improvement (ΔR² < 0.005), indicating that the foundation model embeddings already capture sex-related electrophysiological variation. While this supports the feasibility of embedding-only age prediction, ethnicity-specific calibration may still be warranted for equitable clinical deployment.

#### Training Strategy

Age prediction models were trained exclusively on healthy subjects to establish a normative aging baseline. This design ensures that age deviation in diseased subjects reflects disease-associated cardiac aging rather than confounding by pathological ECG morphology in the training data.

Data were split at the patient level using GroupShuffleSplit (80% train / 20% test) with patient_id as the grouping variable, ensuring that multiple recordings from the same patient never appeared in both train and test sets. A fixed random seed (42) was used throughout for reproducibility.

### ElasticNetCV with Nested Cross-Validation

We used ElasticNetCV with a nested cross-validation architecture. The outer split (GroupShuffleSplit) divided patients into train and test. Within the training set, ElasticNetCV performed hyperparameter selection via 5-fold GroupKFold inner cross-validation, again grouped by patient_id. This nested design prevents information leakage at the patient level during both model selection and evaluation.

The hyperparameter search grid spanned five L1 ratios (0.1, 0.3, 0.5, 0.7, 0.9) and 20 alpha values per ratio, automatically determined by scikit-learn’s alpha path. ElasticNet was chosen over Ridge or Lasso because it combines L1 sparsity (feature selection among 1,024 embedding dimensions) with L2 regularization (stable coefficient estimation in the presence of correlated features), and the cross-validation procedure selects the optimal trade-off data-adaptively.

### Age-Bin Weighting

To counteract the non-uniform age distribution in both datasets (which are skewed toward middle-aged adults), sample weights were computed using inverse-square-root frequency weighting across 5-year age bins. For each age bin *b* with count *n_b_*, the raw weight was computed as:

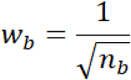

Then, all weights were normalized by dividing by their mean:

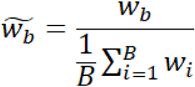

where *B* is the total number of bins. This ensures the average normalized weight equals 1.0. This approach upweights underrepresented age extremes (young and elderly subjects) without the instability of pure inverse-frequency weighting 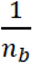, while preserving the overall scale of the loss function.

### Feature Standardization

All features were standardized to zero mean and unit variance using StandardScaler fitted on the training set and applied to the test set, preventing data leakage from test set statistics.

### Age Deviation and Age Acceleration

Two complementary metrics were computed to quantify the discrepancy between predicted and chronological age:

**Age deviation** was defined as the raw residual:

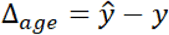

where 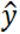 represents the predicted age and *y* denotes the chronological age. This metric is intuitive but inherits the well-known regression-to-the-mean artifact whereby young subjects tend to have positive deviations and old subjects negative deviations.

**Age acceleration** was computed as the residual after regressing predicted age on chronological age using ordinary least squares (*OLS*) fitted on the healthy training set:

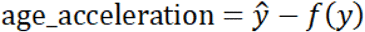

where 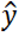 is the predicted age, *y* is the chronological age, and *f*(*y*) represents the linear regression fit. This correction removes the systematic age-dependent bias and isolates the component of predicted age that is independent of chronological age. Age acceleration was used as the primary metric for disease-specific and mortality analyses. The sufficiency of linear OLS correction was validated by comparing linear, quadratic, and spline correction models, which yielded negligible differences in corrected prediction accuracy and near-identical residual slopes (Supplementary Figure S5).

### Disease-Specific Age Acceleration Analysis

For each disease category, age acceleration was compared between affected subjects and the healthy reference group using Welch’s *t*-test (unequal variances assumed). Effect sizes were quantified using Cohen’s d with pooled standard deviation. To account for multiple comparisons across 30 disease categories in PTB-XL (individual SCP codes with *n* ≥ 100) and 12 ICD-10-based disease groups in MIMIC, p-values were corrected using the Benjamini-Hochberg false discovery rate (FDR) procedure at *α=0.05*.

Bootstrap confidence intervals (1,000 iterations with replacement) were computed for the overall diseased-versus-healthy delta and Cohen’s *d* to assess sampling variability and provide robust uncertainty estimates.

### Conduction Bias Analysis

Conduction abnormalities (e.g., bundle branch blocks) produce distinctive ECG morphological changes that may confound embedding-based age prediction, the model may interpret conduction-altered waveforms as age-related changes rather than disease-specific patterns. To investigate this, we implemented a three-configuration sensitivity analysis in PTB-XL:

**Configuration 1 (Embedding only):** 1,024 embedding dimensions + sex. This is the baseline model and reflects the default feature set.
**Configuration 2 (Embedding + Task probabilities):** 1,024 embeddings + 150 ECGFounder task probability outputs + sex (1,175 features total). Task probabilities encode the model’s diagnostic predictions and may capture conduction-specific information that the embedding does not explicitly separate from aging.
**Configuration 3 (Embedding + Disease class):** 1,024 embeddings + sex + one-hot encoded disease labels (top 30 SCP codes with *n ≥ 100*). This configuration directly provides disease identity to the age predictor and serves as an upper bound for conduction bias correction.

For each configuration, a separate ElasticNetCV model was trained on healthy subjects using the identical nested CV procedure. Age deviation was then predicted for all diseased subjects, and the conduction bias was quantified as the Cohen’s *d* between conduction-abnormality patients and non-conduction diseased patients. A reduction in Cohen’s d across configurations would indicate that the confound is attributable to auxiliary features (task probabilities or disease labels) rather than an intrinsic property of the embedding space.

In MIMIC, conduction abnormalities were identified via report-based keyword matching rather than ICD codes, as conduction-specific ICD codes (I44–I45) are infrequently assigned in billing data. The conduction bias was assessed by comparing age acceleration between report-identified conduction patients and the remaining diseased population.

### Lead Configuration Comparison

To evaluate the feasibility of ECG-derived biological age estimation from reduced-lead recordings relevant to wearable and mobile ECG devices, the entire analysis pipeline was independently repeated for each lead configuration (12-lead, 6-lead, 1-lead). Each configuration used its own embedding extraction, ElasticNetCV training on healthy subjects, patient-level split, and age deviation computation.

Performance was compared using *R²* (variance explained), *MAE* (mean absolute error in years), and Cohen’s *d* for disease-versus-healthy discrimination. Sex-stratified analyses were performed for each configuration to assess whether reduced-lead performance varies by sex.

### High-Error Patient Analysis

Patients with absolute age deviation exceeding 15 years were classified as “high-error” cases. Disease enrichment in the high-error group relative to the normal-error group was assessed using Fisher’s exact test with FDR correction. This analysis identifies which diagnostic categories are most strongly associated with large prediction errors, providing insight into which ECG morphologies are most confounded with or most informative about cardiac aging in the embedding space.

### Mortality Analysis (MIMIC-IV-ECG)

To assess the prognostic value of ECG-derived age acceleration, we performed survival analysis on the MIMIC-IV-ECG cohort.

### Survival Data Preparation

To avoid immortal time bias, only the first ECG recording per patient was retained for mortality analysis. Follow-up time was defined as the interval from the first ECG date to either the date of death or the end of the study period (latest recorded date in the dataset), whichever came first. Patients with zero or negative follow-up time were excluded.

### Cox Proportional Hazards Regression

Cox proportional hazards models were used to estimate the association between age acceleration and all-cause mortality, adjusting for chronological age and sex. The proportional hazards assumption was assessed using Schoenfeld residuals. Hazard ratios (HRs) are reported per 1-year and per 5-year increase in age acceleration. Four nested models were compared: (1) age acceleration alone, (2) chronological age alone, (3) chronological age + sex, and (4) the full model (age acceleration + chronological age + sex).

### Concordance Index

Harrell’s concordance index (C-index) was computed for each of the four nested models to quantify discriminative ability. Bootstrap confidence intervals (1,000 iterations) were computed for the difference in C-index between the full model and the age + sex baseline to assess the incremental discriminative value of the ECG-derived biomarker.

### Kaplan-Meier Analysis

Kaplan-Meier survival curves were constructed by dividing patients into quartiles of age acceleration. The log-rank test was used to compare survival between the highest (Q4, most accelerated aging) and lowest (Q1, most decelerated aging) quartiles.

### Landmark Analysis

To verify that the mortality association was not driven by acute peri-recording events (e.g., critically ill patients who received an ECG shortly before death), landmark analyses were performed at 0, 30, 90, and 180 days post-ECG. At each landmark, patients who died before the landmark time were excluded, and the Cox model was re-estimated. Stable hazard ratios across landmarks would support a genuine long-term prognostic association rather than an artifact of acute illness.

### Subgroup Analyses

Subgroup analyses stratified by sex (male, female) and age group (<65, 65–75, ≥75 years) were conducted to identify populations in which ECG-derived age acceleration is most informative for mortality prediction.

### Statistical Methods

All continuous comparisons between groups used Welch’s *t-test* (unequal variances assumed). Multiple comparison correction was performed using the Benjamini-Hochberg FDR procedure throughout. Effect sizes are reported as Cohen’s *d* with 95% bootstrap confidence intervals. Survival analyses used the lifelines Python package. A two-sided *p < 0.05* (after FDR correction where applicable) was considered statistically significant.

### Software and Reproducibility

All analyses were implemented in Python 3.12. Key dependencies include scikit-learn 1.x (ElasticNetCV, StandardScaler, GroupShuffleSplit, GroupKFold), scipy (Welch’s *t*-test, Fisher’s exact test), statsmodels (FDR correction, LOWESS smoothing), lifelines (Cox regression, Kaplan-Meier estimation), and matplotlib (visualization). ECGFounder embeddings were extracted using the publicly available model weights (PKUDigitalHealth/ECGFounder) with the Net1D architecture from hsd1503/resnet1d. All analysis scripts are available at https://github.com/BIMSBbioinfo/ECG_age. A fixed random seed (42) was used throughout to ensure full reproducibility. Computations were performed on the TRUBA high-performance computing cluster (TÜBİTAK ULAKBİM) and the MDC Berlin computing infrastructure.

## Data availability

Core data is available at https://physionet.org/content/mimic-iv-ecg/1.0/ and https://physionet.org/content/ptb-xl/1.0.3/

## Acknowledgements

We thank Akalin lab members for comments on the manuscript. We especially thank Madalin Patrascu for his help deploying the ECGage web app. AA is supported by Helmholtz Validation Fund. The numerical calculations reported in this paper were partially performed at TUBITAK ULAKBIM, High Performance and Grid Computing Center (TRUBA resources).

## Contributions

A.A. conceptualized, planned the project and did initially feasibility assessment and analysis. D.A. with the help of F.G. jointly executed all of the computational analyses. D.A made all the figures, completed benchmarks and wrote the initial draft of the manuscript. A.S. built the user-friendly app with the help of D.A. D.A., A.A., A.S. and F.G. edited the manuscript. A.A. acquired funding and supervised the project.

