## Supplementary material for "ECG-derived age deviation predicts cardiovascular diseases across lead configurations and cohorts"

**Supplementary Materials**

**Supplementary Table S1.** Landmark analysis of ECG-derived age acceleration and all-cause mortality in MIMIC-IV-ECG.

| **Landmark (days)** | **n patients** | **n events** | **HR per year** | **95% CI** | **p-value** | **C-index** |
| --- | --- | --- | --- | --- | --- | --- |
| 0 | 160,493 | 26,902 | 1.005 | 1.004–1.006 | 1.88×10⁻¹⁷ | 0.746 |
| 30 | 156,477 | 22,886 | 1.007 | 1.006–1.008 | 2.02×10⁻²⁸ | 0.744 |
| 90 | 153,725 | 20,134 | 1.009 | 1.007–1.010 | 3.18×10⁻³⁶ | 0.741 |
| 180 | 151,428 | 17,837 | 1.010 | 1.009–1.011 | 6.78×10⁻⁴² | 0.739 |

**Supplementary Table S2.** Concordance index comparison across nested Cox regression models.

| **Model** | **C-index** |
| --- | --- |
| **Age acceleration only** | 0.510 |
| **Chronological age only** | 0.743 |
| **Age + sex** | 0.745 |
| **Full (accel + age + sex)** | 0.746 (95% CI: 0.744–0.748) |

**Supplementary Table S3.** SCP-ECG code descriptions. SCP-ECG diagnostic code abbreviations and descriptions used in the PTB-XL analysis. All codes and descriptions are from the PTB-XL dataset's official SCP-ECG annotation standard.

| **SCP Code** | **SCP-ECG Statement Description** |
| --- | --- |
| NORM | Normal ECG |
| DIG | Digitalis-effect |
| ISCAL | Ischemic in anterolateral leads |
| LNGQT | Long QT-interval |
| ISC_ | Ischemic ST-T changes (non-specific) |
| ISCLA | Ischemic in lateral leads |
| NDT | Non-diagnostic T abnormalities |
| NST_ | Non-specific ST changes |
| IMI | Inferior myocardial infarction |
| LVH | Left ventricular hypertrophy |
| INJAL | Subendocardial injury in anterolateral leads |
| CLBBB | Complete left bundle branch block |
| CRBBB | Complete right bundle branch block |
| IRBBB | Incomplete right bundle branch block |
| 1AVB | First degree AV block |
| LAFB | Left anterior fascicular block |
| LPFB | Left posterior fascicular block |
| IVCD | Non-specific intraventricular conduction disturbance |
| AFIB | Atrial fibrillation |
| PVC | Ventricular premature complex |
| PACE | Normal functioning artificial pacemaker |
| RVH | Right ventricular hypertrophy |

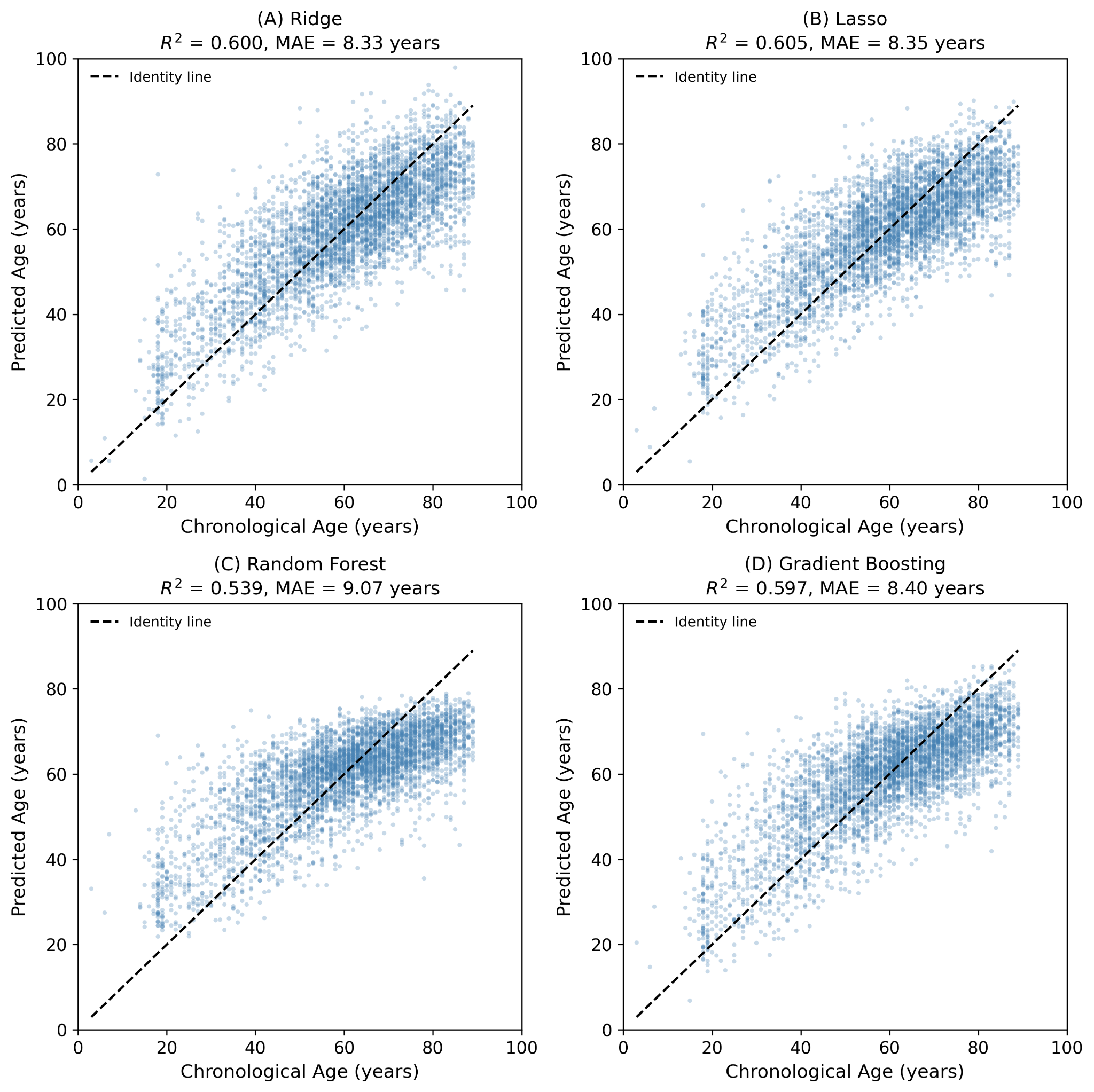

**Fig. S1:** Model comparison (Ridge/Lasso/GB/RF)Age prediction performance across four regression models on the full PTB-XL dataset. Scatter plots of predicted versus chronological age for (A) Ridge regression (R²=0.600, MAE=8.33 years), (B) Lasso regression (R²=0.605, MAE=8.35 years), (C) Random Forest (R²=0.539, MAE=9.07 years), and (D) Gradient Boosting (R²=0.597, MAE=8.40 years). Dashed lines represent perfect prediction (y=x). All models were trained on the full dataset (healthy + diseased) for model selection purposes; the primary analysis used ElasticNetCV trained on healthy subjects only (R²=0.611, MAE=8.31 years). Linear models performed comparably to ensemble methods, supporting the use of ElasticNetCV for subsequent analyses.

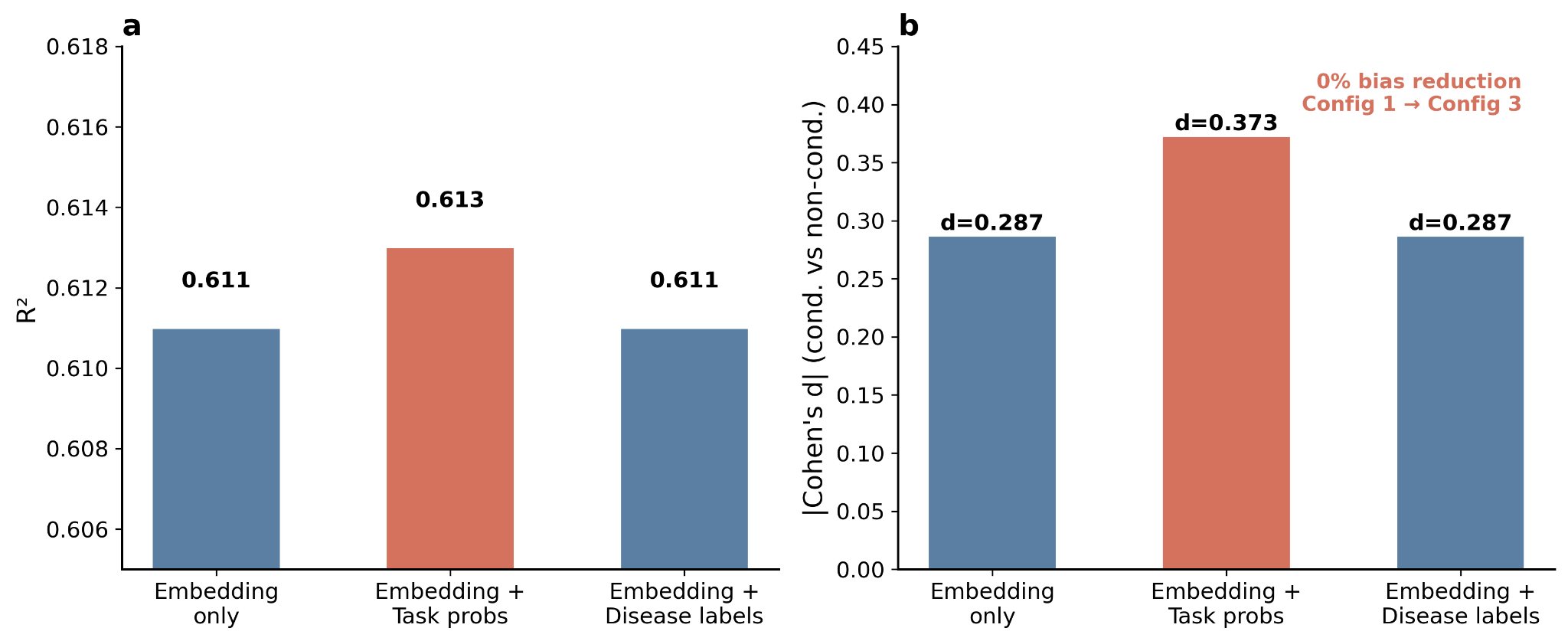

**Fig. S2:** Conduction 3-config sensitivity. Three feature configurations tested to determine whether the conduction bias is correctable: Configuration 1 (embedding only, d=−0.287), Configuration 2 (embedding + 150 task probabilities, d=−0.373), and Configuration 3 (embedding + 30 disease one-hot labels, d=−0.287). The 0% bias reduction from Configuration 1 to 3 indicates that the conduction bias is intrinsic to the ECGFounder embedding space. Configuration 2 amplified the bias, suggesting task probability outputs encode conduction-specific information that further separates conduction from aging signals.

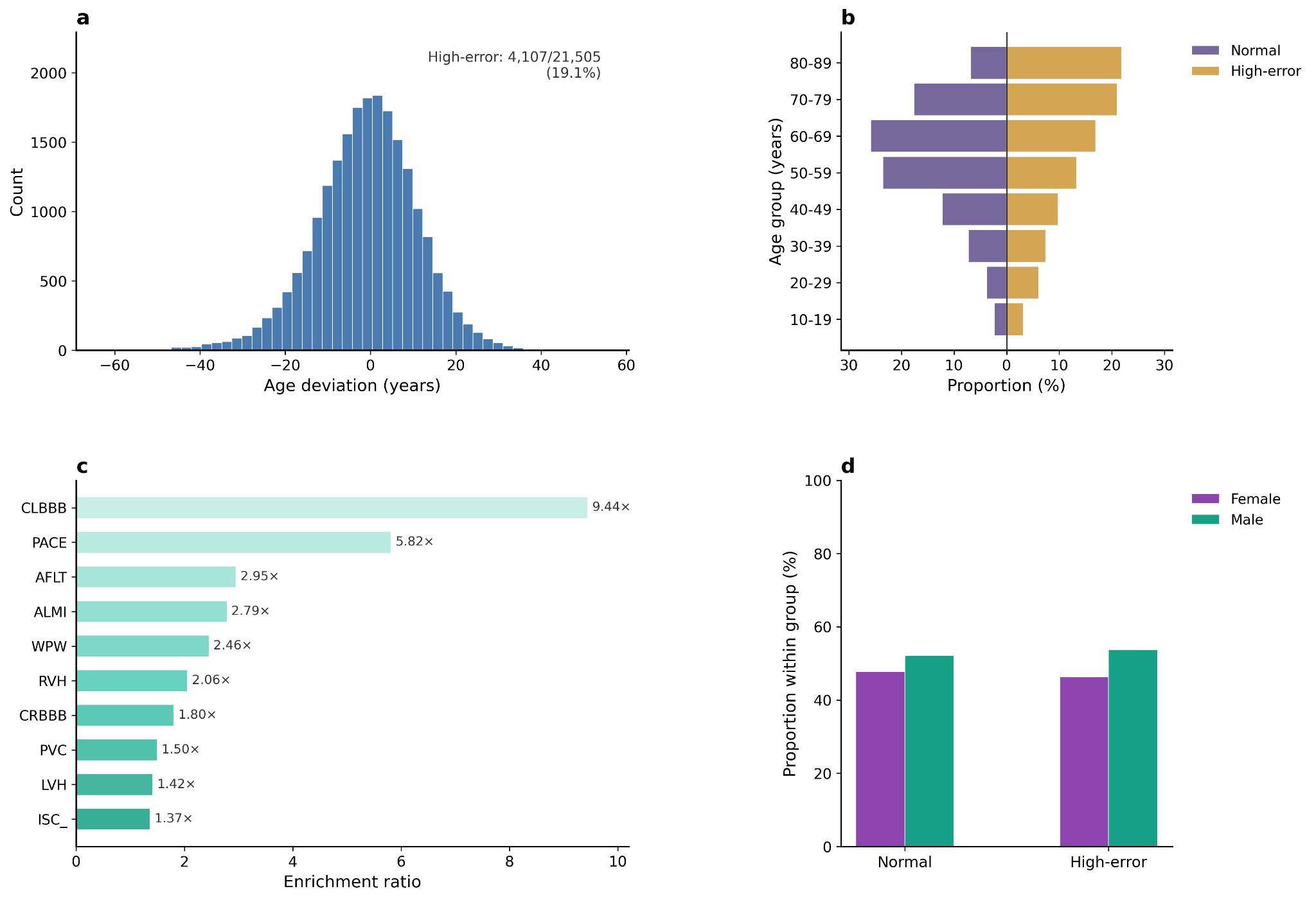

**Figure S3. High-error patient analysis in the PTB-XL cohort** High-error patient analysis in PTB-XL. (a) Distribution of age deviation across all recordings. (b) Age distribution comparison between normal-error and high-error patients. (c) Disease enrichment ratios in the high-error group relative to normal-error, showing that complete left bundle branch block (CLBBB, 9.44×) and pacemaker rhythm (PACE, 5.82×) are the most over-represented conditions. (d) Sex distribution comparison between normal-error and high-error groups.

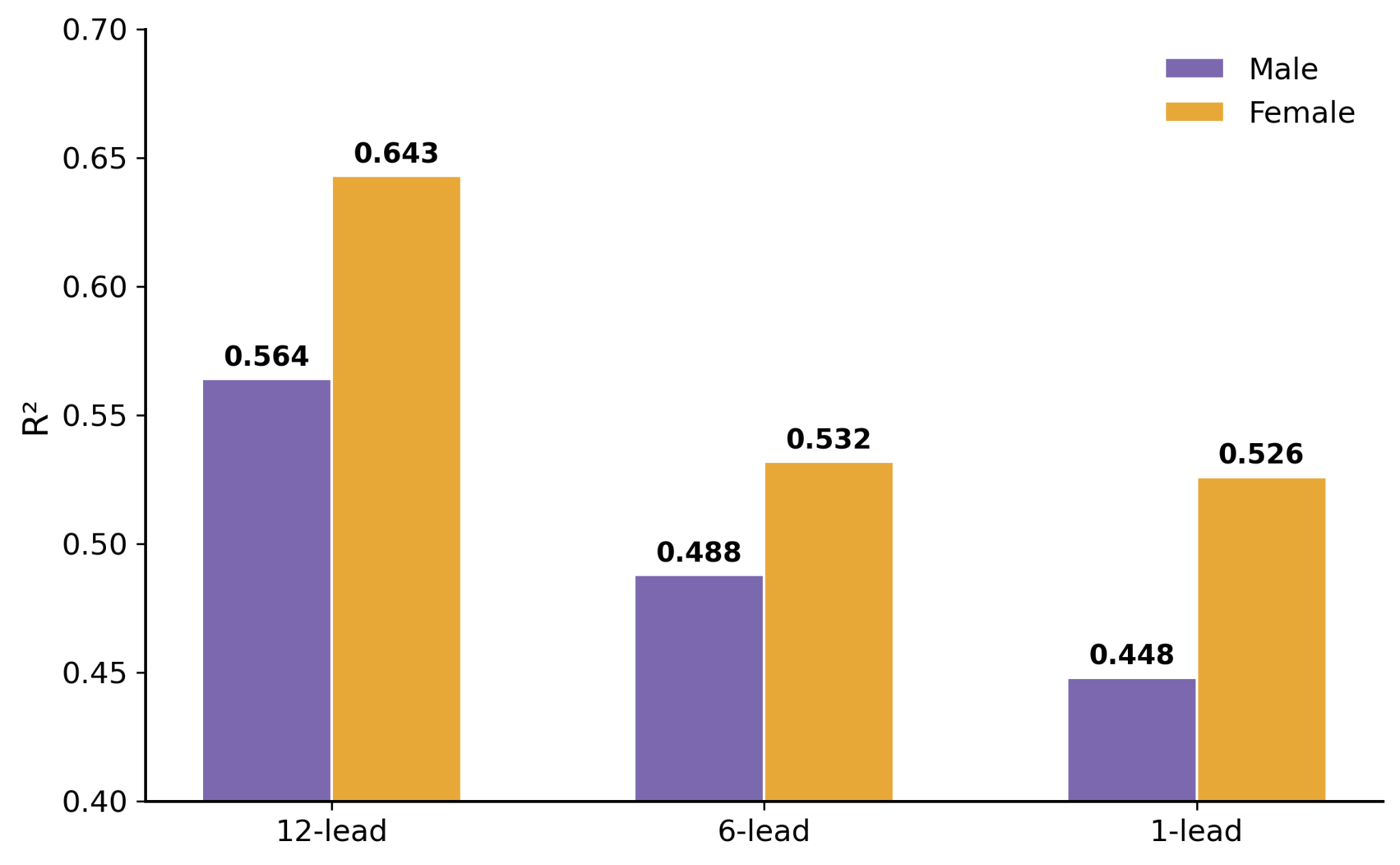

**Fig. S4:** Sex-stratified lead comparison. Age prediction performance stratified by sex across all three lead configurations. Females consistently showed higher R² (12-lead: 0.643 vs 0.564; 6-lead: 0.532 vs 0.488; 1-lead: 0.526 vs 0.448), suggesting a more consistent aging signal in female ECG embeddings.

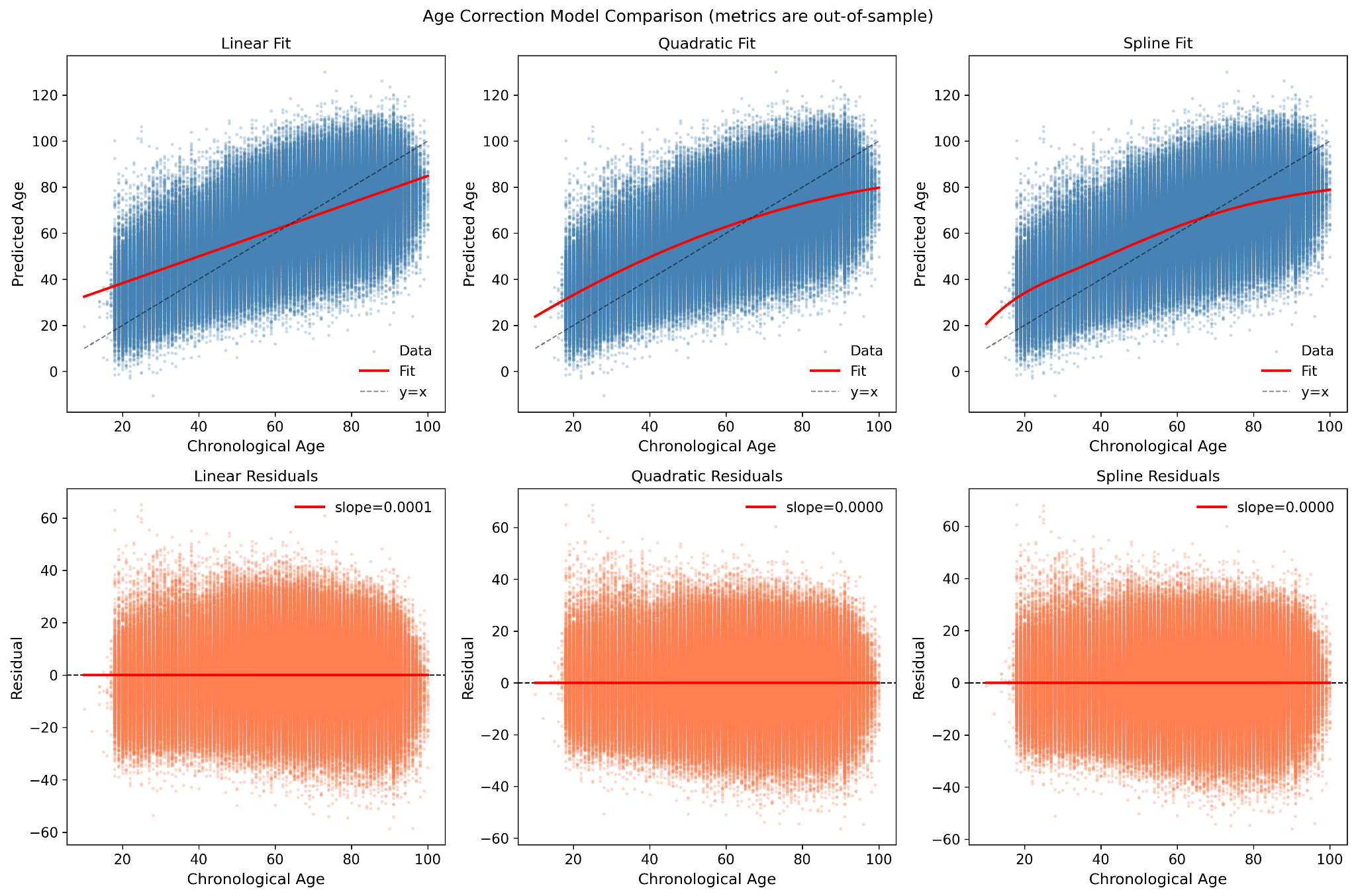

**Figure S5.** Comparison of linear, quadratic, and spline age correction models in MIMIC-IV-ECG. Top row: predicted versus chronological age with fitted correction curves. Bottom row: residual plots after correction, showing near-zero slopes across all three methods (linear: 0.0001, quadratic: 0.0000, spline: 0.0000), confirming the sufficiency of linear OLS correction.
